# Genome admixture analysis of 1,030 Ugandan infants with neonatal sepsis and hydrocephalus demonstrates geographical stratification of population disease risk

**DOI:** 10.64898/2026.03.16.26348489

**Authors:** Mercedeh Movassagh, Lucinda Newbury, Christine Hehnly, Andrew J. Whalen, Mallory Peterson, Enrique Mondragon-Estrada, Jessica E. Ericson, Jasmine Smith, Misaki Sasanami, Davis Natukwatsa, John Mugamba, Peter Ssenyonga, Justin Onen, Kathy Burgoine, Lijun Zhang, Peter Olupot-Olupot, Elias Kumbakumba, Emmanuel Wegoye, Moses Ochora, Ronald Mulondo, Edith Mbabazi-Kabachelori, Claudio Fronterre, James Broach, Joseph N. Paulson, Sarah U. Morton, Steven J. Schiff

**Affiliations:** Department of Neurosurgery, Yale University School of Medicine, New Haven, CT; Harvard TH Chan School of Public Health, Department of Biostatistics, Boston, MA; Dana Farber Cancer Institute, Department of Data Science, Boston, MA; Boston Children’s Hospital, Division of Newborn Medicine, Department of Pediatrics, Boston, MA; University of Birmingham, Birmingham, UK; Boston Children’s Hospital and Harvard Medical School, Department of Pathology, Boston, MA; Penn State College of Medicine, Department of Neurosurgery, Hershey, PA; Penn State College of Medicine, Department of Pediatrics, Hershey, PA; Penn State College of Medicine, Department of Biochemistry and Molecular Biology and Institute for Personalized Medicine, Hershey, PA; Cure Hospital, Mbale, Uganda; Mulago National Referral Hospital, Mulago, Uganda; Mbale Regional Referral Hospital, Mbale, Uganda; Case Western Reserve University, Department of Population and Quantitative Health Sciences, School of Medicine, Cleveland, Ohio, USA; Mbarara University of Science & Technology (MUST), Mbarara, Uganda; Department of Paediatrics and Child Health, Soroti University; Department of Data Science, N-Power Medicine, Inc., Redwood City, CA, USA; Department of Pediatrics, Harvard Medical School, Boston, MA; Department of Epidemiology of Microbial Diseases, Yale University School of Public Health, New Haven, CT

**Keywords:** Genetic Admixture, Genome Sequencing, Hydrocephalus, Neonatal Sepsis, Population genetics, Uganda

## Abstract

**Background:** Neonatal disorders such as post-infectious hydrocephalus exhibit a higher incidence in Africa, where the intricate relationships between genetic ancestry, environmental exposures, and other risk factors likely contribute to the increased incidence.

**Methods:** To start to characterize the common genetic architecture of Ugandan infants, we analyzed genome sequencing data from 1,030 Ugandan infants recruited from studies targeting neonatal sepsis and hydrocephalus. We employed genetic admixture analysis and integrated geospatial data to examine the relationships between genetic backgrounds and disease prevalence within this cohort.

**Results:** Our results identified four distinct genetic admixture groups, each correlating strongly with specific geographic distributions across Uganda. Notably, a predominance of one admixture group, most common in northern Uganda, was overrepresented in the participants with post-infectious hydrocephalus.

**Conclusion:** This study underscores the importance of genetic factors in disease manifestation at the population level, and a role for such precision public health approaches in complex neonatal disorders in African populations.

## Background

Disease susceptibility varies across populations due to genetic and environmental factors, and studies have found the greatest diversity of genetic variants among African populations. However, current genomic studies in African peoples are often underpowered to identify genetic risk markers because of the dearth of broad African representation^1,2^. The Global Biobank Meta-analysis Initiative, a recent effort to assess genetic contribution to disease risk across diverse cohorts, has identified hundreds of novel coding variants associated with common diseases such as asthma and stroke^3^. Furthermore, genetic ancestry groups underrepresented in reference databases have lower rates of pathogenic variant identification, exacerbating disparities in diagnosis and treatment^4,5^. As a consequence, the accuracy of polygenic disease risk scores remains lower for underrepresented ancestries in research cohorts, highlighting the need to recruit cohorts who reflect our diverse world^6^.

The history of African ancestry is complex, marked by migrations that occurred over thousands of years and aligns with ethnolinguistic divergence on the continent^7^. Genetic evidence from ancient samples identifies substantial agricultural expansions including the most extensive event that coincided with the migration of the Bantu peoples originating in northwest Africa and spreading predominantly southward, followed by an eastward and northward movement^8–10^. A secondary agricultural expansion involved the Nilo-Saharan Africans, who migrated southward into central and eastern Africa^11,12^. Positioned at the western edge of the Horn of Africa, Uganda is an ideal geographical location to study the impact of ancient migrations on African genomics. In the investigation by Gurdasani et al., genotyping and whole genome sequencing techniques reported up to 18 admixture groups within one district within southwest Uganda^13^. Such localized sampling would be unable to demonstrate associations with more extensive geographical coordinates. Recent East African genetic studies are yielding increasing clarity for genetic risk profiling of African peoples^14,15^. An analysis of genetic substructure in southwestern Uganda revealed diverging polygenic risk score variability as compared to Sardinian and Icelandic cohorts in over 30 traits^13,16,17^. The Neuropsychiatric Genetics of African Population-Psychosis (NeuroGAP-Psychosis) demonstrated the utility of low-coverage genome sequencing to detect novel variants^18^. Reports from Uganda indicate a high incidence of neonatal conditions such as hydrocephalus and sepsis, with rates among the highest globally^19–21^. The estimated incidence of infant hydrocephalus in sub-Saharan Africa is 750 per 100,000 births, compared to 68 per 100,000 births in the United States and Canada^20^. A large proportion of hydrocephalus in Uganda is attributed to post-infectious causes, often secondary to neonatal sepsis (NS)^15,19,20^.

In Uganda and sub-Saharan Africa in general, infant hydrocephalus is the most common indication for neurosurgery in early childhood^22^. Genetic risk has been estimated to contribute to 10–20% of congenital hydrocephalus cases in cohorts primarily of European ancestry^23^; however, this proportion is likely to differ in other populations due to differing environmental exposures, infectious burdens, and underlying genetic architectures. In Uganda nearly 60% of infant hydrocephalus is due to a central nervous system infection, often originating from NS, and is termed post-infectious hydrocephalus (PIH)^24–28^. We hypothesize that the increased prevalence of neonatal disorders such as sepsis leading to PIH is due in part to common variants present in the Ugandan population. As susceptibility to infection and associated outcomes have been shown to be influenced by genetic factors in other settings^29,30^, identifying genetic risk can inform more effective strategies to mitigate severe infections in Ugandan infants. As a first step towards this goal, we sought to characterize the primary genetic admixtures present among Ugandan infants. We performed genome sequencing on blood samples collected from participants geographically distributed throughout Uganda and characterized their genetic ancestry and disease correlation. The proportions of four genetic admixture groups correlated with the geospatial location of their residence. Finally, we assessed the correlation of admixture groups among participants with NS and infant hydrocephalus phenotypes.

## Results

### Identification of common variants

Genome sequencing was performed on blood samples from 1,185 infants, including 392 participants from a study of hydrocephalus - encompassing congenital hydrocephalus (CH; excluding spina bifida with myelomeningocele) or post-infectious hydrocephalus (PIH) - and 793 participants from a study of NS^26,31,32^. Participants with hydrocephalus were recruited at the CURE Children’s Hospital of Uganda, a neurosurgical specialty care center in Mbale. Participants with NS were recruited from regional referral hospitals in Mbale (eastern Uganda) and Mbarara (western Uganda; Table 1).

**Table 1.** Cohort description. Abbreviations: SD, standard deviation.

|  |  | *Post Infectious*<br>*Hydrocephalus*<br>*(PIH)* | *Congenital*<br>*Hydrocephalus*<br>*(CH)* | *Neonatal Sepsis*<br>*(NS)* |
| --- | --- | --- | --- | --- |
| **Total** | **N** | **196** | **172** | **662** |
| **Region** | **Eastern N (%)** | **134 (68%)** | **57 (33%)** | **372 (56%)** |
|  | **Western N (%)** | **3 (2%)** | **17 (10%)** | **372 (40%)** |
|  | **Northern N (%)** | **46 (23%)** | **25 (15%)** | **1 (0%)** |
|  | **Central N (%)** | **13 (7%)** | **73 (42%)** | **21 (3%)** |
| **Sex** | **Male N (%)** | **112 (57%)** | **98 (57%)** | **373 (56%)** |
| **Hospital** | **CURE N (%)** | **196 (100%)** | **172 (100%)** | **0 (0%)** |
|  | **Mbarara N (%)** | **0 (0%)** | **0 (0%)** | **374 (56%)** |
|  | **Mbale N (%)** | **0 (0%)** | **0 (0%)** | **288 (43%)** |

After joint variant calling and quality control, including genotype missingness filtering, minor allele frequency (MAF) >0.1 filtering, and exclusion of low-confidence variants, we retained 1,030 genomes with 46,414,185 single nucleotide polymorphisms (SNPs) for downstream analysis. The final analysis cohort included 368 infants with hydrocephalus (n=172 CH; n=196 PIH) and 662 infants with NS (Figure 1A). Identity-by-descent (IBD) analysis was performed using PLINK^33^ and identified no close genetic relationships among individuals, with a calculated coefficient of relatedness near zero.

**Figure 1.**
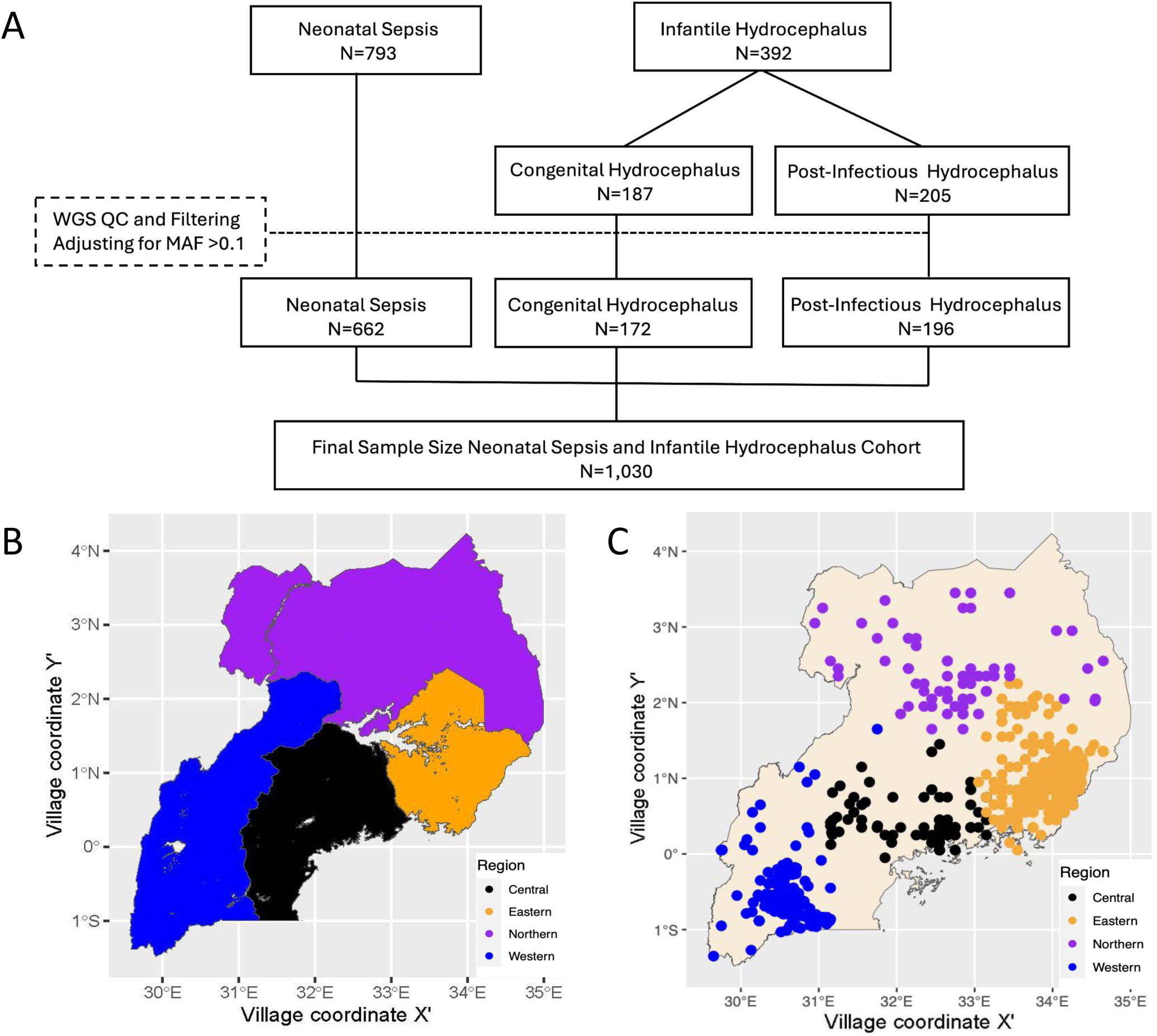
(A) CONSORT Diagram for participant enrollment, sample collection availability, and data processing for each participant diagnosis group. Abbreviations: MAF, minor allele frequency; QC, quality control; and WGS, whole genome sequencing. (B) Map of geographical regions of Uganda. (C) Geospatial distribution of study participants, with each dot indicating an individual after geomasking. Color corresponds to geographical regions.

### Characterizing the genetic structure of the Ugandan cohort through admixture analysis

Uganda is organized into four regions by the Ugandan National Planning Authority. By combining the hydrocephalus and NS cohorts, we achieved substantial representation of individuals from all four described regions in the country (Figure 1B-C). We adopted a statistical framework to estimate genetic admixture. We hypothesized that due to historical migration patterns, individuals would harbor components of multiple local ancestries ^32,33,34,35^. ADMIXTURE^34^ analysis was performed on the combined hydrocephalus and NS cohorts to model individual ancestry proportions with cross-validation error minimized at four ancestry groups (K=4; Figure 2A-B, Supplemental Figure 1). Participants were categorized into genetic Groups 1-4 based on their dominant (largest admixture proportion) ancestral group. There was a strong association between predominant ancestral group and geographical region of residence (chi-square p-value < 2.2×10⁻¹□, Figure 2C-D, Table 2). For example, most participants from the northern region (67/72) were assigned to Group 3, while Group 4 was more common in the western region (88/292). Participants from the central, eastern, and western regions were predominantly classified into Group 1 and Group 2. Overall, individual ancestry assignments and regional residence showed significant correlation (chi-square p-value < 2.2×10⁻¹□ ; Supplemental Figure 2).

**Figure 2.**
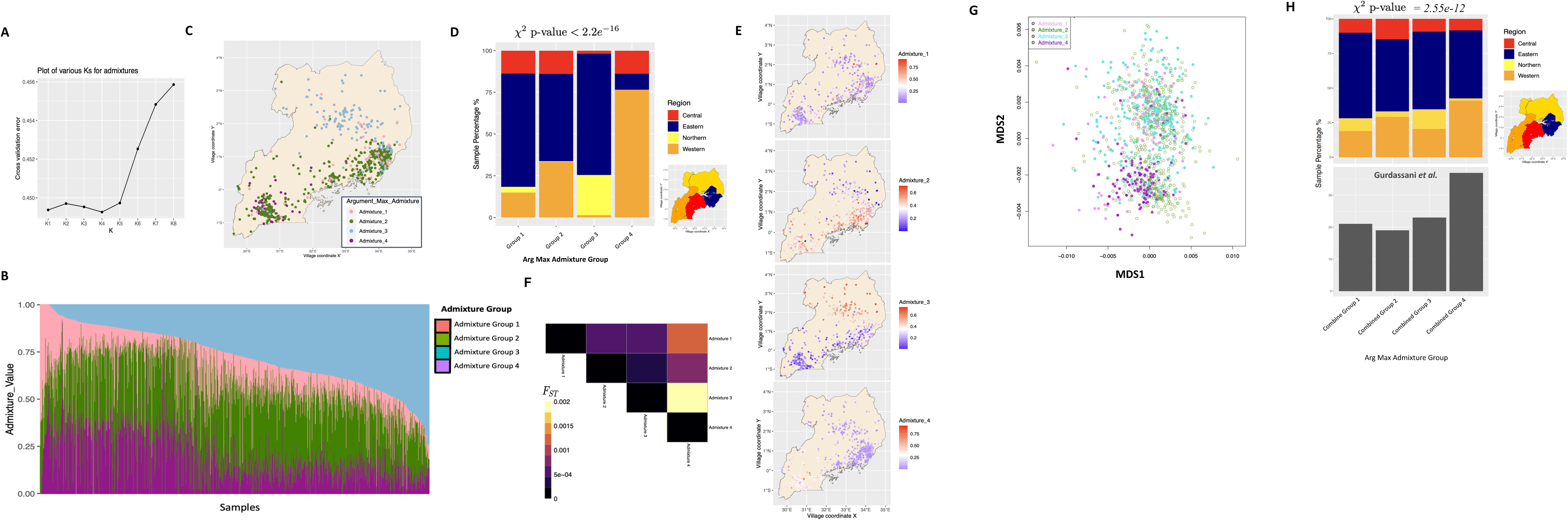
Admixture Analysis Reveals Distinct Ancestral Genetic Groups in 1,030 Ugandan Samples. (A) Optimal number of clusters based on minimizing the cross-validation error of admixture composition for our dataset comprising 1,030 Ugandan samples. In this case, K=4 (K4) emerges as the optimal cluster number, as it yields the lowest cross-validation error among the range of possibilities (K1-K8). (B) Admixture bar plot representing each individual’s composition of the four distinct admixture groups identified in the dataset, with each group denoted by a unique color. (C) Color-coded representation of individuals based on the predominant genetic ancestry, subsequently mapped onto the geographical regions of Uganda according to their residence. Each data point represents a participant, with colors indicating the cluster assigned through K-means clustering. (D) Bar plot showing the distribution of reported residential regions assigned to each predominant ancestral group. (E) Geospatial maps illustrating the proportions of ancestral groups across different regions. The blended colors on the maps signify the proportion of genetic ancestral groups within the participants. (F) Heatmap illustrates the genetic distances between admixture groups based on the highest proportion of admixture assignments (F_ST_ values). Colors in the heatmap correspond to the degree of genetic distance between admixture groups.

**Table 2.** Home region or cohort and predominant genetic admixture. Number of participants from each region (top) or cohort (bottom) based on predominant genetic admixture group (columns).

|  | ADMIXTURE 1 | ADMIXTURE 2 | ADMIXTURE 3 | ADMIXTURE 4 |
| --- | --- | --- | --- | --- |
| **Region** |  |  |  |  |
| **Central** | **12** | **77** | **5** | **16** |
| **Eastern** | **59** | **287** | **203** | **11** |
| **Northern** | **3** | **2** | **67** | **0** |
| **Western** | **13** | **187** | **4** | **88** |
| **Cohort** |  |  |  |  |
| **Neonatal Sepsis** | **30** | **301** | **131** | **88** |
| **Congenital**<br>**Hydrocephalus**<br>**(CH)** | **14** | **62** | **32** | **4** |
| **Post infectious**<br>**hydrocephalus**<br>**(PIH)** | **19** | **90** | **74** | **5** |

Mapping the geographic distribution of ancestry proportions revealed region-specific patterns (Figure 2E). Group 2 and Group 4 ancestry proportions were enriched in the central and western regions, while Group 3 ancestry was concentrated in the north. Group 1 proportions were relatively lower overall, with some clustering in the central and eastern regions.

To assess genetic differentiation between inferred ancestral groups, we estimated pairwise fixation indices (F_ST_) using PLINK^33^. Mean F_ST_ values were modest, consistent with fine-scale population structure, with the highest differentiation observed between Group 3 and Group 4 (mean F_ST_ = 0.002), followed by Group 1 and Group 4 (mean F_ST_ = 0.0012), and Group 2 and Group 4 (mean F_ST_ = 0.0007; Figure 2F).

### Inclusion of published cohort in admixture analysis

#### Integration of External Ugandan Cohort

To assess the reproducibility of the previously defined admixture groups, we incorporated published low-coverage genome sequencing data from 364 healthy adults residing in a sub-county of southwestern Uganda^13^. After genotype merging and harmonization, 26,605,619 variants were found in both datasets, representing approximately 42% of the 46,414,185 variants used in the original admixture analysis. Filtering to variants with a minor allele frequency (MAF) greater than 0.1 retained 14,875,173 variants for downstream analysis.

As with the original analysis prior to including the published cohort, ADMIXTURE modeling identified four clusters (K=4) as the optimal solution, minimizing cross-validation error (0.10). Those clusters were called Combined Groups 1-4. No phasing or permutation was applied during dataset integration, a limitation we acknowledge; however, the admixture group assignments remained stable despite potential batch differences.

#### Stability of Admixture Groups

In the combined cohorts, the previously identified Combined Group 3 continued to be the most common among individuals from the northern region (Supplemental Figure 3A-B). Most participants from the western region were assigned to Combined Group 4 (38%) and Combined Group 2 (27%), mirroring the distributions observed in the initial analysis prior to including the published cohort.

Among the external samples from the Gurdasani study, 37% were classified into Combined Group 4 and 19% into Combined Group 2, consistent with the regional assignment of western Ugandan ancestry. These findings support the reproducibility and robustness of the four inferred admixture groups, although small differences between datasets are expected given differences in geographic focus, and technical protocols.

### Association of local genetic ancestral group with disease state

#### Association of Ancestry Groups with Hydrocephalus Subtypes

Within the hydrocephalus cohort, we assessed whether ancestry groups were differentially associated with CH versus PIH status. A multivariate Kruskal-Wallis test revealed a significant difference in ancestry group distributions across hydrocephalus subtypes (p = 2.51×10⁻□, η² = 0.10). Post-hoc Wilcoxon tests indicated that admixture Group 1 was significantly associated with CH (p = 0.03, η² = 0.11), Group 3 was significantly associated with PIH (p = 6.89×10⁻□, η² = 0.22), and Group 4 was significantly associated with CH (p = 1.03×10⁻□, η² = 0.25) (Figure 3A–D). No significant association was observed between Group 2 and hydrocephalus subtype.

**Figure 3.**
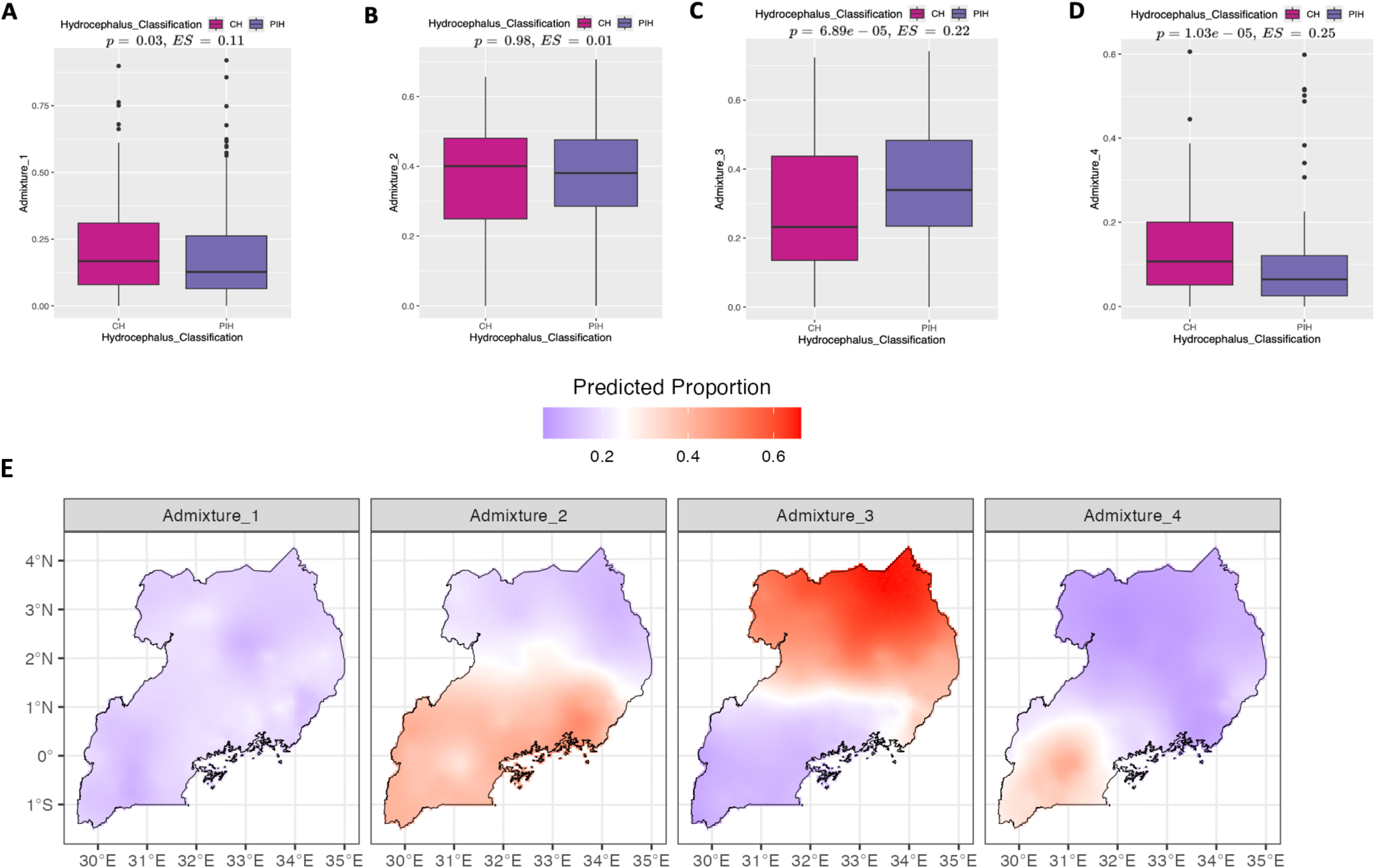
Admixture Group Associations with Disease Phenotypes. (A-D) Box plot depicting the Ratio Admixture Associations among congenital hydrocephalus (CH), and postinfectious hydrocephalus (PIH) groups. Squared correlation ratio represents the effect size (ES). (E) Geospatial Alignment of Genetic Admixture within Uganda. Geostatistical modeling of the proportions for each of the four identified admixture clusters, minimizing mean squared error for each group, resulting in separate maps. Map colors indicate predicted distribution of each admixture group within the country as calculated by log-odds transform of the Gaussian geospatial model.

These findings suggest that ancestral genetic backgrounds represented by Group 1 and Group 4 may be associated with an increased likelihood of congenital hydrocephalus, while Group 3 ancestry may be associated with an increased likelihood of post-infectious hydrocephalus. However, given the observational nature of the study, these associations should not be interpreted as evidence of causality.

### Geospatial prediction of local genetic ancestral groups

#### Geostatistical Modeling of Ancestry in Uganda

We employed a geostatistical modeling technique that accounts for spatial correlation across a continuous geographic surface, to minimize the impact of artificial boundaries and provide continuous spatial correlation for non-neighboring locations despite close physical proximity^35,36^. Specifically, we individually fit spatial Gaussian process models to the proportions of each of the four identified genetic ancestry groups. Model parameters were selected to minimize mean squared prediction error. This continuous approach enables smooth forecasting of ancestral group distributions across Uganda, including in unobserved locations, without imposing artificial boundaries (Figure 3E).

Spatial prediction maps derived from this model revealed patterns broadly consistent with known ancient migration and ethnolinguistic histories within Uganda^37^. Although no direct validation is possible, the alignment of predicted genetic distributions with established historical trends supports the relevance of our ancestry groups to regional population movements.

## Discussion

### Genetic Structure and Diversity Within Uganda

Disease susceptibility arises from a complex interplay of genetic and environmental factors. Our studies highlight the extensive genetic diversity within Uganda, a country shaped by millennia of human migration across sub-Saharan Africa. By sequencing and analyzing a geographically distributed cohort of 1,030 Ugandan infants, we identified four major ancestral groups corresponding to distinct regions of the country. We generated smooth spatial maps from these proportional admixtures that were found to reflect known ancestral migration patterns into this region of central Africa.

Our admixture modeling and spatial analyses revealed notable regional genetic differentiation, with the largest genetic distances observed between groups corresponding geographically to northern and western Uganda. The magnitude of genetic differentiation within Uganda, exemplified by an F_ST_ value of 0.002 between Admixture Groups 3 and 4, is comparable to that observed between continental ancestry groups in large-scale biobank studies. These findings underscore the remarkable genetic substructure present within a single African country and are consistent with previous observations of substantial fine-scale structure in neighboring populations, such as those in Kenya ^11,42^.

### Genetic Associations with Infant Hydrocephalus

Within the hydrocephalus cohort, we observed associations between specific ancestry groups and hydrocephalus subtypes. Admixture Group 1 and Group 4 were associated with congenital hydrocephalus (CH), while Group 3 was associated with post-infectious hydrocephalus (PIH). While these observations suggest possible contributions of ancestry to disease susceptibility, it is important to emphasize that our study design cannot establish causality, and environmental factors may also contribute to the observed differences.

### Limitations

Despite its strengths, our study has several important limitations. Additional information regarding regional rates of neonatal sepsis and hydrocephalus is needed to better interpret the contribution of genetic risk to these outcomes. Although relatively large for a genome-sequencing effort in Uganda, the sample size remains modest for ancestry-disease association analyses, limiting our ability to detect more granular genetic differences. The use of low-coverage sequencing introduces potential inaccuracies in variant calling and reduces sensitivity for detecting rare variants. Recruitment from hospital-based cohorts may introduce biases compared to the general population, particularly for the NS cohort, which was drawn from two distinct hospitals in eastern and western Uganda. Although we observed robust admixture groupings even when integrating an external dataset without phasing or permutation, future studies should aim to incorporate larger, geographically dispersed cohorts and higher-coverage sequencing data to characterize fine-scale population structure more accurately.

## Conclusion

In conclusion, this study demonstrates the presence of distinct regional genomic ancestries within the modern Ugandan population. By leveraging admixture analysis, spatial modeling, and disease phenotype data, we provide insights into the rich genetic structure of Uganda and its potential relevance to understanding disease risk. These findings highlight the critical importance of incorporating genomic diversity into global health efforts and may inform future precision public health strategies aimed at improving disease prevention and treatment across diverse populations.

## Methods

### Study Enrollment

To study the genomes of the Ugandan cohorts, ethical oversight was provided by the Mbarara University of Science and Technology Research Ethics Committee, the CURE Children’s Hospital of Uganda Institutional Review Board, the Pennsylvania State University and Yale University Institutional Review Boards, and the Ugandan National Council on Science and Technology. The participants were recruited as previously described as part of studies of infant hydrocephalus and neonatal sepsis in Uganda^26,31,32^, and collected prior to the current Genome Data Sharing policy by the US National Institutes of Health. Specimens were shared with US institutions under Material Transfer Agreements and US Centers for Disease Control permits. A Genome Data Sharing Policy was screened by NIH and approved by all the above ethics and government oversight committees which, in brief, allowed sharing the results of genomic analysis only in aggregate so that individual participants could not be re-identified, and that no public deposition of genomic sequences be shared. The data sharing agreement also stipulated that participant location information was reported only with a precision of 0.1 degrees (equivalent to 11 x 11 km at the equator), and that participants from grid locations with populations of less than 500 people were not mapped.

The hydrocephalus cohort^26,32^ recruitment was conducted at the CURE Children’s Hospital of Uganda, a specialized pediatric neurosurgical hospital in eastern Uganda. This hospital serves as a referral center for children with hydrocephalus nationwide. Mothers were required to be at least 18 years old and capable of providing written informed consent in a local language. Infants were eligible to participate in the study if they were 90 days of age or younger and met specific criteria for PIH or CH. To be included in the PIH group, infants had to weigh more than 2.5 kilograms, have no history of hydrocephalus at birth, and either a history of febrile illness and/or seizures preceding the onset of clinically apparent hydrocephalus or alternative findings on imaging or endoscopy indicative of prior cerebral ventriculitis. For inclusion in the CH group, infants also had to weigh more than 2.5 kilograms and display non-infectious causes of hydrocephalus on computed tomography (CT) scan or during endoscopy, such as obstruction of the Aqueduct of Sylvius, hemorrhage as the cause of hydrocephalus, or congenital malformations of the nervous system. Exclusion criteria included prior nervous system surgery or evidence of nervous system communication with the skin. Infants likely to have a genetic syndrome with anomalies other than hydrocephalus were excluded. Spina bifida associated myelomeningocele, which commonly leads to hydrocephalus, was excluded from this CH cohort.

The neonatal sepsis cohort^26,31^ included term neonates weighing more than 2 kilograms who were recruited with clinical neonatal sepsis at the Mbarara Regional Referral Hospital in Western Uganda and the Mbale Regional Referral Hospital in Eastern Uganda. Mothers were required to be at least 18 years old and capable of providing informed written consent in a local language. Neonatal sepsis was clinically defined as the presence of one of the following three combinations of signs: (1) axillary temperature >37·5°C, lethargy, and poor feeding, (2) axillary temperature <35·5°C, lethargy, and poor feeding, or (3) full fontanelle or seizures, axillary temperature >37·5°C, and poor feeding. Neonates were excluded from recruitment if they had major congenital abnormalities, had a history of perinatal asphyxia, or had received antibiotics for more than 24 hours before recruitment^26,31^. Fifteen infants in the sepsis cohort later developed hydrocephalus, of which 8 were not analyzed and 7 were reassigned to the hydrocephalus cohort.

### Sample Collection and Preservation

Blood samples from infants were collected as previously described using DNA/RNA Shield (Zymo, CA, USA) and frozen at −80°C or placed in liquid nitrogen^32^.

### DNA Extraction and Whole Genome Sequencing

Nucleic acid extraction was done as previously described with 500 μL of blood processed using the ZymoBIOMICS DNA Miniprep Kit (Zymo, CA, USA) in accordance with the manufacturer’s protocols with the addition of bead lysis with two sized zinc oxide beads^32^. Whole genome sequencing was done at the New York Genome Center with 500 ng of high molecular weight DNA using the Illumina TruSeq Nano kit and sequenced to 4-6x coverage.

### Sample alignment and preprocessing

Alignment, preprocessing, and subsequent downstream analysis followed published pipelines (Supplemental Figure 4). In brief, FASTQ files were obtained from approximately 5x low coverage sequencing and the reads were aligned to the human genome (GRCh38) using *Burrower Wheelers Algorithm* (BWA)^38^. The binary alignment map (BAM) files were sorted utilizing *SAMtools*^39^, and the duplicated reads marked using *Picard* tools^40^. All samples were jointly called using *Genomic Analysis ToolKit* (GATK)^41^ HaplotypeCaller in GVCF mode and merged using GenomicsDBImport prior to final variant calling. dbSNP138 was used for compatibility with GATK’s base quality recalibration model trained on historical datasets; we acknowledge that newer versions (e.g., dbSNP154) offer broader coverage. We employed the GATK Gaussian mixture model utilizing the GATK Variant/base-recalibrator function for variant calling. The model was trained using reference datasets, including hapmap version 3.3. for hg38, 1000G_omni2.5 for hg38, and the 1000G_phase1 SNPs dataset, all based on *Homo sapiens* assembly 38, in conjunction with dbsnp138. Several filtering criteria were applied to ensure the quality of the variant calls, including read coverage, quality by depth, root mean square of the mapping quality of reads across all samples, rank sum test for mapping quality of reference versus alternate (ALT) base reads, rank sum test for relative positioning of REF vs ALT alleles within reads, strand bias estimated using Fisher’s exact t-test (reverse vs forward strand), strand bias estimated using odds ratio (reverse vs forward strand), and depth of allele per sample. A tranches cutoff of 99.5 was employed to refine the selection of variants. Coefficient of relatedness was calculated for the cohort using KING v2.2.5^42^ and no pairs exceeded a Pi_HAT of 0.125.

### Common variant set

For downstream analysis, we used PLINK^33^ for variant filtration and association studies using a linkage-disequilibrium pruned SNP set (--indep-pairwise 50 5 0.2 in PLINK). To ensure the reliability of our results, particularly due to the relatively low sequencing depth, we applied stringent filtering criteria^43^: variant MAF >0.1, which reduced variants from 1,685,806,711 to 46,414,185 single nucleotide polymorphisms. Then 155 samples with >5% genotype missingness were removed.

### Admixture analysis

Admixture analysis was conducted using the ADMIXTURE software^34^ on genotype data that passed quality control filtering. We evaluated a range of potential cluster numbers (K=2 to K=10) to determine the optimal model, selecting K=4 based on minimization of cross-validation error (Supplemental Figure 1). Each participant was assigned a dominant ancestry group, defined as the ancestry component with the largest proportional contribution. These groups were labeled Groups 1 through 4. We assessed the relationship between dominant ancestry group and region of residence using a chi-square test, which demonstrated a highly significant association (chi-square p-value < 2.2×10⁻¹□). To further characterize genetic differentiation among inferred ancestral groups, we calculated pairwise fixation indices (FST) using the Weir and Cockerham estimator implemented in PLINK^33^.

### Reproducibility of Admixtures

To evaluate the reproducibility of inferred admixture groups, we integrated low-coverage genome sequencing data from 364 adults residing in a sub-county in southwestern Uganda, a random subset of the cohort previously described by Gurdasani et al^13^. For the 364 samples, 30,679,188 sites were identified, including 1,761,023 multiallelic sites. Variant call format files from the external cohort were lifted from human reference genome GRCh37 (b37/hg19) to GRCh38 (hg38) using LiftoverVcf in Picard^40^ and then merged with the internal dataset using bcftools^39^. Only variants present in both datasets were retained. After merging, 26,605,619 variants were common across both datasets, representing approximately 42% of the 46,414,185 variants used in the initial analysis. Variants with a minor allele frequency greater than 0.1 and genotype missingness <5% were selected for downstream analyses, resulting in a final set of 14,875,173 SNPs for admixture modeling. ADMIXTURE analysis was rerun on the combined dataset. No genotype phasing or imputation was performed prior to merging datasets, and no permutation testing was applied to account for batch effects between studies. This decision reflects typical challenges in integrating independently sequenced cohorts but allows for assessment of general cluster stability.

### Geospatial Categorization and Mapping of Regions

In all figures, the geomasked coordinates (latitude and longitude) correspond to the centroid of the 0.1-degree grid squares enclosing the participants’ resident villages, with additive random draws from a normal distribution with 0.05 degrees standard deviation to both protect privacy of location and to enhance visibility of overlapping cases from the same grid square, unless stated otherwise. Maps of Ugandan administrative areas were obtained from the National Planning Authority and Uganda Bureau of Statistics containing the regional divisions of the country as well as district, county, sub county, parish, and village boundaries. All country shapefiles for Uganda were projected into World Geodetic System 1984 (WGS84) coordinates in all figures. Participant case data was labeled with the administrative area information by spatial joins in ArcMap (ArcGIS Desktop 10.8.2, Esri, Redlands, CA, USA) and *R* v.4.3.0^44^.

### Geospatial Prediction Estimation

For geospatial estimation, we assumed that the log-odds transform, 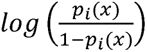 of the proportion, *p_i_* (*x*), of ancestral group *i* at location *x* can be expressed as:

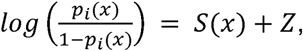

where *S(x)* is a stationary Gaussian spatial process with an isotropic Matérn covariance function, and *Z* are independent zero-mean Gaussian variables ^40^. For each location *x* in a *5km*^2^ grid, we predicted the expected proportion, *E*[*p_i_* (*x*)] of each ancestral group, *i* ∈ {1,2,3,4}, for an individual living at that location. We aimed to enforce the constraint that the ancestral group proportions for a given location *x* sum to 1, ie. ∑*_i_* (*E*(*p_i_* (*x*))=1. To do so, we replaced the prediction for Group1, which displayed as the least accurate in terms of mean squared error for the observed locations, with the difference:

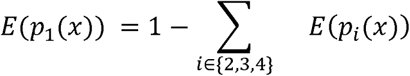

between 1 and the sum of the proportions of the remaining three ancestral group proportions.

### Statistics

Wilcoxon rank sum tests were used to compare continuous variables, Kruskal-Wallis test were used to compare proportions, while chi-square and ANOVA tests were used to compare categorical traits. Significant thresholds were determined using Bonferroni correction for number of tests, and unadjusted p-values were reported.

## Data Availability

Raw genomic data were not consented for open access. The downstream analysis files as well as code for statistical, geospatial and anthropometric analysis and plots are present at the data folder on github.

github.com:Mercedeh66/WGS_Uganda.git

## List of abbreviations

CH: Congenital Hydrocephalus
*F_ST_*: Fixation Index
GS: Genome Sequencing
PIH: Post-infectious hydrocephalus
MAF: Minor allele frequency
NS: Neonatal Sepsis
SNVs: Single nucleotide variants
VCF: Variant call file

## Declarations

### Data Availability

The downstream analysis files are present at the data folder github.com:Mercedeh66/WGS_Uganda.git.

### Code Availability

The code for statistical, geospatial and anthropometric analysis and plots can be found at github.com:Mercedeh66/WGS_Uganda.git.

### Ethics declarations and competing interests

M.M. and E.M-E. performed the whole-genome sequencing (WGS) analysis, and M.M. and J.N.P. conducted the statistical analysis. M.M., and L.Z. handled the preprocessing of the WGS data. L.H., C.F., M.M., A.W. and M.S. performed the geo-spatial analysis and mapping. C.H. and J.S. were responsible for sample extraction and library preparation. S.U.M., M.M., S.J.S., R.M., J.N.P., and J.B. designed the project, and M.M., S.U.M., S.J.S., L.H., C.H., J.N.P. and M.P. contributed to the writing of the article. K.B., S.J.S., C.H., A.W., P.S., J.M., J.O., E.K., E.W., K.N., and P.O.O. were involved in sample collection, and S.J.S, M.P., E.M., R.M., J.E. and D.N. collected and reviewed the clinical and phenotypic data associated with the project and did the study enrollment.

### Consent for publication

All authors have read and reviewed the paper, and there are no conflicts of interest to declare.

## Acknowledgment

We would like to thank Dr. Rafael Irizarry and the Irizarry lab for the helpful discussions on the topic. We thank Deepti Gurdasani for helpful discussions on their dataset.

