## Supplemental Figure 1, Supplemental Figure 2, Supplemental Figure 3, Supplemental Figure 4 for "Genome admixture analysis of 1,030 Ugandan infants with neonatal sepsis and hydrocephalus demonstrates geographical stratification of population disease risk"

**Supplemental Figures**

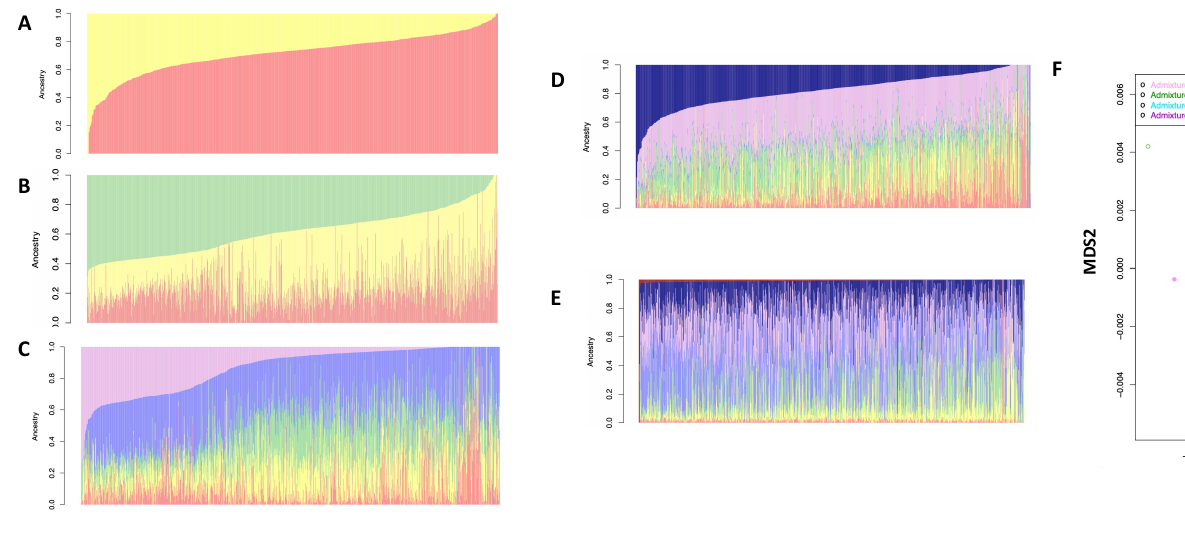
**Supplemental Figure 1.** ADMIXTURE bar plot representing each individual’s composition of the admixture groups varying from K=1 (A) to K=5 (E).

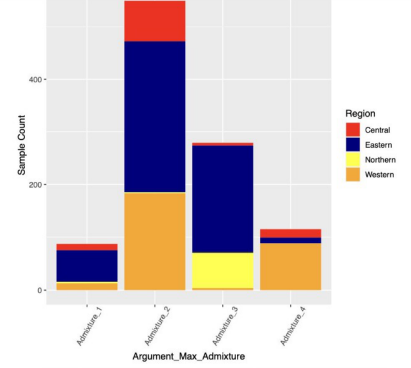

**Supplemental Figure 2.** Association between dominant ancestry group and region of residence by total participant count.

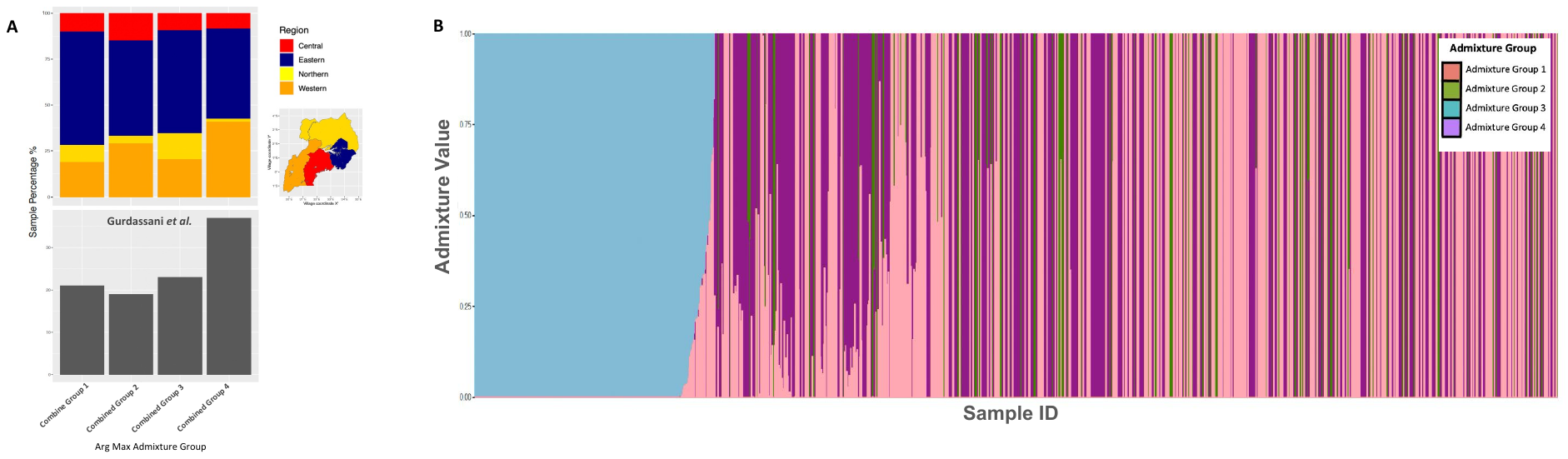
**Supplemental Figure 3.** (A) Bar plot of Admixture assignments for the Ugandan infant cohort when combined with the Gurdasani et al.^13^ low depth genome sequencing data (above). Proportion of the Gurdasani et al. dataset assigned to each Admixture group (below). (B) Admixture bar plot representing each individual proportion of the four distinct admixture groups identified in the combined dataset, with each group denoted by a unique color.

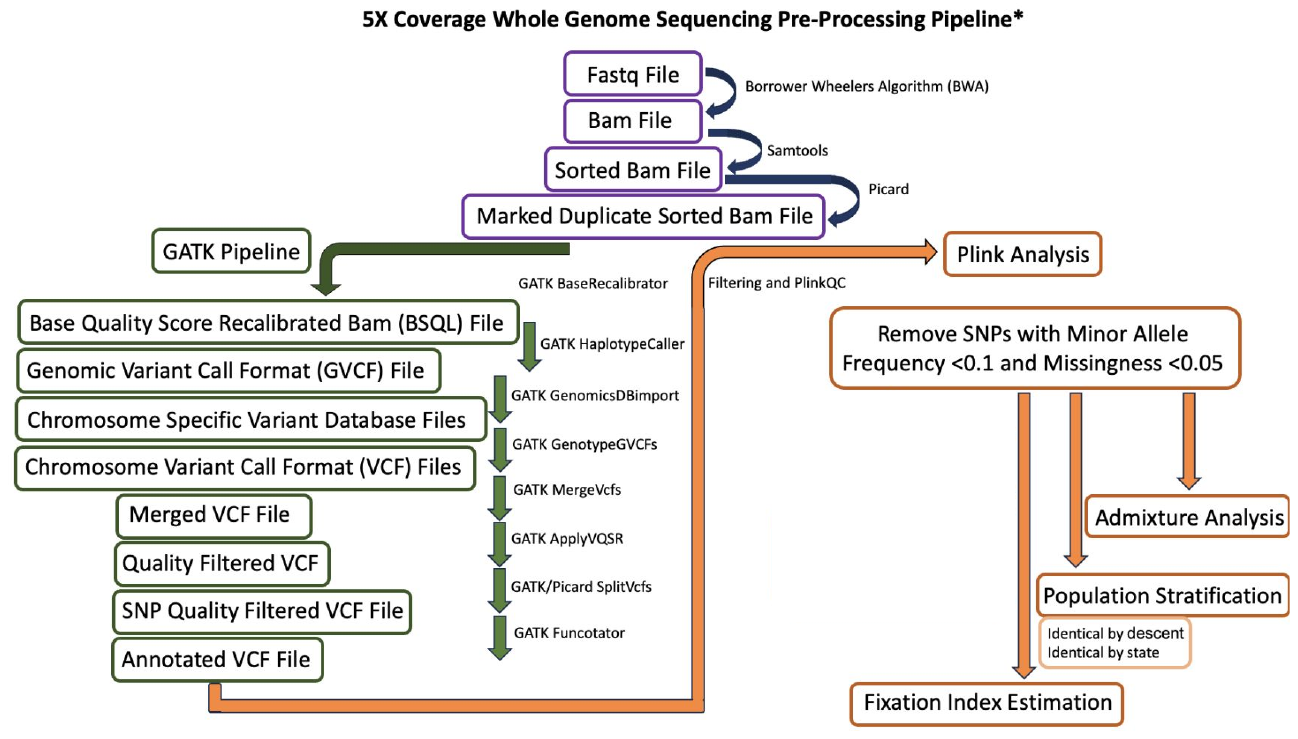
**Supplemental Figure 4.** Genetic data processing pipeline.
